# Obesity Prediction in Young Adults from the Jerusalem Perinatal Study: Contribution of Polygenic Risk and Early Life Exposures

**DOI:** 10.1101/2023.09.05.23295076

**Authors:** Hagit Hochner, Rachely Butterman, Ido Margaliot, Yechiel Friedlander, Michal Linial

## Abstract

We assessed whether adding early life exposures to a model based on polygenic risk score (PRS) improves prediction of obesity risk. We used a birth cohort with data at birth and BMI and waist circumference (WC) measured at age 32. The PRS was composed of SNPs identified in GWAS for BMI. Linear and logistic models were used to explore associations with obesity-related phenotypes. Improvement in prediction was assessed using measures of model discrimination (AUC), and net reclassification improvement (NRI). One SD change in PRS was associated with a significant increase in BMI and WC. These associations were slightly attenuated (13.7%-14.2%) with the addition of early life exposures to the model. Also, higher maternal pre-pregnancy BMI was associated with increase in offspring BMI and WC (p<0.001). For prediction obesity (BMI ≥ 30), the addition of early life exposures to the PRS model significantly increase the AUC from 0.69 to 0.73. At an obesity risk threshold of 15%, the addition of early life exposures to the PRS model provided a significant improvement in reclassification of obesity (NRI, 0.147; 95% CI 0.068-0.225). We conclude that inclusion of early life exposures to a model based on PRS improves obesity risk prediction in an Israeli population-sample.

## Introduction

Obesity is a worldwide major health problem. A large body of literature has established a clear link between obesity with adverse health outcomes, including diabetes mellitus, coronary heart disease, stroke, heart failure and increased mortality (1, 2).

Risk factors for obesity have been studied extensively, and in general, they are divided into: demographic and socio-economic (e.g., education, and socioeconomic status) (3, 4); environmental (e.g., unhealthy diet, smoking, and sedentary lifestyle), (5) and genetic factors (6, 7). More recently, large-scale genome-wide association studies (GWASs) and meta-analyses have identified approximately 200 loci that are associated with adiposity traits across different populations worldwide (8, 9). However, SNPs identified in GWASs explain little of obesity variance (7, 10, 11). Combining these SNPs into a polygenic risk score (PRS) has proven valuable, offering a quantifiable measure of inherited predisposition toward a trait, such as BMI (12). Developing and validating obesity-specific PRS represents a breakthrough in personalized obesity and obesity-related disease risk prognosis (13). Obesity-related PRS have been constructed in adults and children, displaying significant associations with numerous phenotypes across diverse populations (14, 15).

Beyond the aforementioned risk factors, current research indicates that fetal and early-life characteristics (like birth weight) play an important role in determining childhood and adulthood obesity. Maternal overnutrition, reflected in part by greater maternal pre-pregnancy body mass index (mppBMI) and gestational weight gain (GWG), has been consistently linked with offspring adiposity across their lifespan (16). Other maternal attributes (e.g., educational attainment and maternal smoking during pregnancy) have also emerged as risk factors (17, 18).

Early Intervention to prevent childhood and young adulthood obesity is key in attenuating obesity epidemic (19). Predictive models gauge the likelihood of specific diseases, such as obesity. While risk prediction based on a single predictive factor have limited prognostic accuracy, combining multiple factors into a prediction model, enhances accuracy (20).

This study’s aim was to construct validated prediction models of young-adulthood obesity using BMI-related PRS, socio-demographic characteristics, and early-life traits. These attributes were collected through the Jerusalem Perinatal Study (JPS), alongside follow-up measurements of obesity-related traits at ages 32-35. This study assesses whether the collected data at early life can enhance the prediction of obesity at adulthood, as projected by PRS.

## Methods

The JPS encompasses a subset of 17,003 births in Jerusalem from 1974 to 1976 (21). Data consist of demographic and sociodemographic information, medical conditions of the mother during current and previous pregnancies, and offspring birth weight from birth certificates or maternity ward logbooks. In addition, and following delivery, postpartum interviews regarding lifestyle and maternal health provided data on gestational age, smoking status, height and pre-pregnancy weight, end of pregnancy weight and gynecological history.

The JPS Follow-up study conducted between 2007 and 2009 includes a sample of about 1,400 offspring from the original cohort (22). Sampling frame included singletons and term (gestational age ≥36 weeks) births without congenital malformations. We obtained a stratified sample of eligible individuals, where the strata were defined by mppBMI and birth weight. Both low (≤2500 grams) and high (≥4000 grams) birth weight as well as mothers with obesity (BMI ≥27) were over-sampled. Detailed information on data collection has been previously described (22). Height, weight, waist measurements and fasting blood samples were collected following standard protocols.

Genomic DNA was extracted using the salting-out method, then genotyping using the Affymetrix Biobank array (comprising of 587,021 variants). We used standard exclusion criteria for the genetic variants and individual samples (23). PLINK 2.0 (24) is employed for variant and individual filtering, resulting in 398,491 variants and 872 offspring for this study. Imputation was done using a combined panel of the 1000 genomes (25) and The

Ashkenazi Genome Consortium (*n* = 128) as reference (26). Imputed variants with MAF ≥1% and quality score

≥0.9 are retained (hard calls), generating a high-quality genetic dataset with nearly 7M genetic variants per individual available for subsequent analyses.

### Study variables

The primary outcome examined was offspring’s BMI and waist circumference (WC) at age 32. BMI and WC, treated as continuous variables. Additionally, this study also examined two outcomes: (i) obesity, defined as BMI ≥30, and (ii) central obesity, defined as WC > 8^th^ decile of the gender specific WC distribution.

PRS was computed using parameters derived from BMI-related GWAS. The association study employed BMI summary statistics from various European ancestry populations, encompassing 339,224 individuals, and a LD reference panel of 503 European samples from 1000 Genomes Phase 3, Ver 5 (25, 27). This panel consists of 2,100,302 common variants (MAF ≥1%). The PRS was established by the LDPred computational algorithm, a Bayesian method that calculates a posterior mean effect for all variants using external weights, then shrinks them based on linkage disequilibrium (28).

Principle component analysis (PCA) was applied on genotyped samples of both parents and offspring samples (n=2,575) along with Jewish reference panel (n=174) using PC-Air (29). Rather than assigning offspring to a single ethnicity, the first five PCs were included as covariates in regression models to account for variations in ancestry within the Jewish population in Israel.

In addition, the following explanatory variables were examined: socioeconomic status (SES) based on father’s occupation (grouped into six categories: 1 lower class - 6 upper class), mother’s years of education (continuous), parental age at offspring’s birth (continuous), birth weight (continuous), gestational age (weeks, continuous), mppBMI (continuous) and GWG (kg, continuous), and maternal smoking during pregnancy (categorical: current smoker vs. never smoked or smoked in the past).

### Statistical analyses

Linear regression models were used to explore the associations of PRS, socio-demographic and perinatal characteristics, independent of each other, with BMI and WC. Four sets of models were constructed. Model 1 included sex as covariate, Model 2 included PRS adjusted for ethnicity (PC1-PC5) and sex, in Model 3 integrated socio-demographic characteristics, and Model 4 incorporated prenatal characteristics. Since these models were nested it enabled an assessment of each variable group’s contribution to explained BMI and WC variability. For categorical outcomes (obesity and central obesity), we used logistic regression analyses. Since the analyses of our data set used probability weights, the likelihood-based tests may not be valid. An alternative method is comparing predicted probabilities from each model to assess their performance.

The area under the curve (AUC) was applied in evaluating the effectiveness of various diagnostic-models in separating between subjects with and without obesity (30). Internal validity of the prediction model was assessed by 10-fold cross-validation.

The Net reclassification improvement (NRI) index (31), was used to compare the saturated model (Model 4) and the restricted model (Model 2) for their ability to classify each subject either with or without obesity. The overall NRI which is the sum of improvement in reclassification of each obesity group was examined by χ2 values. A category-free version of the NRI was also presented (32) to ascertain risk increases to any extent for subjects with obesity under the saturated model (Model 4) compared to the restricted model (Model 2), and similarly whether risk decreases to any degree for subjects without obesity.

This study was approved by the Institutional Review Board of the Hadassah-Hebrew University Medical Center and all participants provided informed consent.

## Results

**Table 1** presents maternal and offspring birth characteristics, offspring PRS and obesity-related traits at age 32. Among mothers (around age 27.5), mppBMI was 23.8 kg/m^2^ and GWG was 11.3 kg. Mean offspring BMI at age 32 was 26.7, with greater WC among males than females. Overweight prevalence was higher in males (41.1%) than females (25.9%).

**Table 1.**
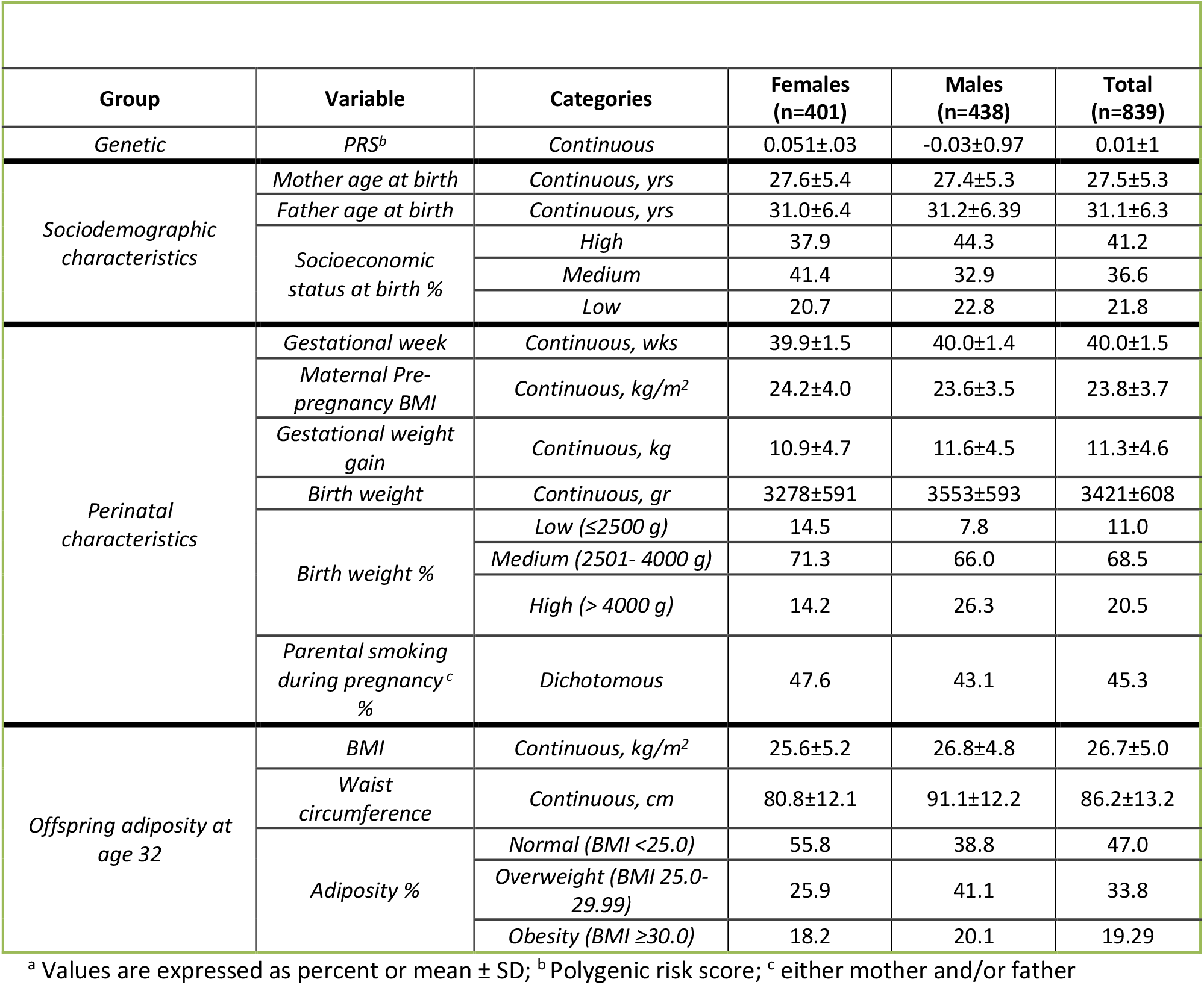
Descriptive characteristics of study participants by sex groups.

Linear regression models’ outcomes are displayed in **Table 2a**, examining the genetic, socio-demographic, and prenatal factors associated with offspring BMI. In Model 2, PRS showed a significant average BMI increase (1.4 kg/m^2^) per one standard deviation. Notably, this PRS association is independent of sex and the first 5 PCs. Socio-demographic contributed minimally to the explained variability of offspring BMI (ΔR^2^=0.0058; p=0.183; Model 3).

**Table 2a.** Parameter Estimates from **Linear** Regression models of BMI as predicted by various groups of Risk Factors.

| Group | Variable | Model 1 |  | Model 2 |  | Model 3 |  | Model 4 |  |
| --- | --- | --- | --- | --- | --- | --- | --- | --- | --- |
|  |  | β | (95% CI) | β | (95% CI) | β | (95% CI) | β | (95% CI) |
| 1 | Sex | 1.74*** | 0.86 - 2.61 | 1.68*** | 0.85 - 2.51 | 1.60*** | 0.77 - 2.44 | 1.70*** | 0.87 - 2.53 |
|  | PRS |  |  | 1.40*** | 0.97 - 1.83 | 1.41*** | 0.99 - 1.84 | 1.21*** | 0.77 - 1.65 |
|  | PC1 |  |  | 0.70** | 0.23 - 1.17 | 0.89*** | 0.36 - 1.41 | 0.73** | 0.20 - 1.27 |
| 2 | PC2 |  |  | 0.20 | -0.20 - 0.61 | 0.13 | -0.29 - 0.56 | 0.06 | -0.36 - 0.47 |
|  | PC3 |  |  | -0.04 | -0.56 - 0.48 | -0.01 | -0.52 - 0.51 | 0.05 | -0.46 - 0.56 |
|  | PC4 |  |  | 0.30 | -0.26 - 0.86 | 0.35 | -0.21 - 0.90 | 0.37 | -0.17 - 0.91 |
|  | PC5 |  |  | 0.36* | 0.06 - 0.66 | 0.37* | 0.05 - 0.69 | 0.26 | -0.08 - 0.60 |
|  | Paternal SES |  |  |  |  | 0.19 | -0.10 - 0.48 | 0.11 | -0.17 - 0.39 |
| 3 | Maternal age |  |  |  |  | -0.07 | -0.19 - 0.06 | -0.08 | -0.20 - 0.04 |
|  | Paternal age |  |  |  |  | 0.03 | -0.08 - 0.14 | 0.02 | -0.09 - 0.12 |
|  | Birth weight |  |  |  |  |  |  | 0.19 | -0.46 - 0.84 |
|  | Gestational week |  |  |  |  |  |  | -0.23 | -0.56 - 0.09 |
| 4 | ppmBMI |  |  |  |  |  |  | 0.39*** | 0.29 - 0.50 |
|  | Parental smoking |  |  |  |  |  |  | 0.6 | -0.25 - 1.44 |
|  | GWG |  |  |  |  |  |  | 0.07 | -0.02 - 0.17 |
|  | R<sup>2</sup> | 0.0346 |  | 0.1271 |  | 0.1329 |  | 0.1964 |  |

PRS - polygenic risk score, PC – principal component SES – Socioeconomic status; ppmBMI – pre-pregnancy maternal BMI; GWG – gestational weight gain; \* p<0.05; \*\* p<0.01; \*\*\* p<0.001

By incorporating perinatal variables to the model, the explained variability of offspring BMI increased significantly (R^2^ rose from 0.133 (Model 3) to 0.196 (Model 4), F_5, 823_ = 12.9; p<=0.001). Each SD change in mppBMI was associated with 0.39 kg/m^2^ increase in offspring BMI (p≤0.001). The association between PRS and offspring BMI was slightly attenuated (though remained significant) upon the addition of perinatal variables into the model.

**Figure 1a** illustrates observed and adjusted offspring BMI means across PRS deciles, showing substantial shifts between the 3 lower and 3 upper deciles. For example, mean observed and predicted BMI levels for offspring located at the upper PRS decile were nearly 5 kg/m^2^ higher compared with BMI mean levels in the lowest decile of PRS.

**Figure 1.**
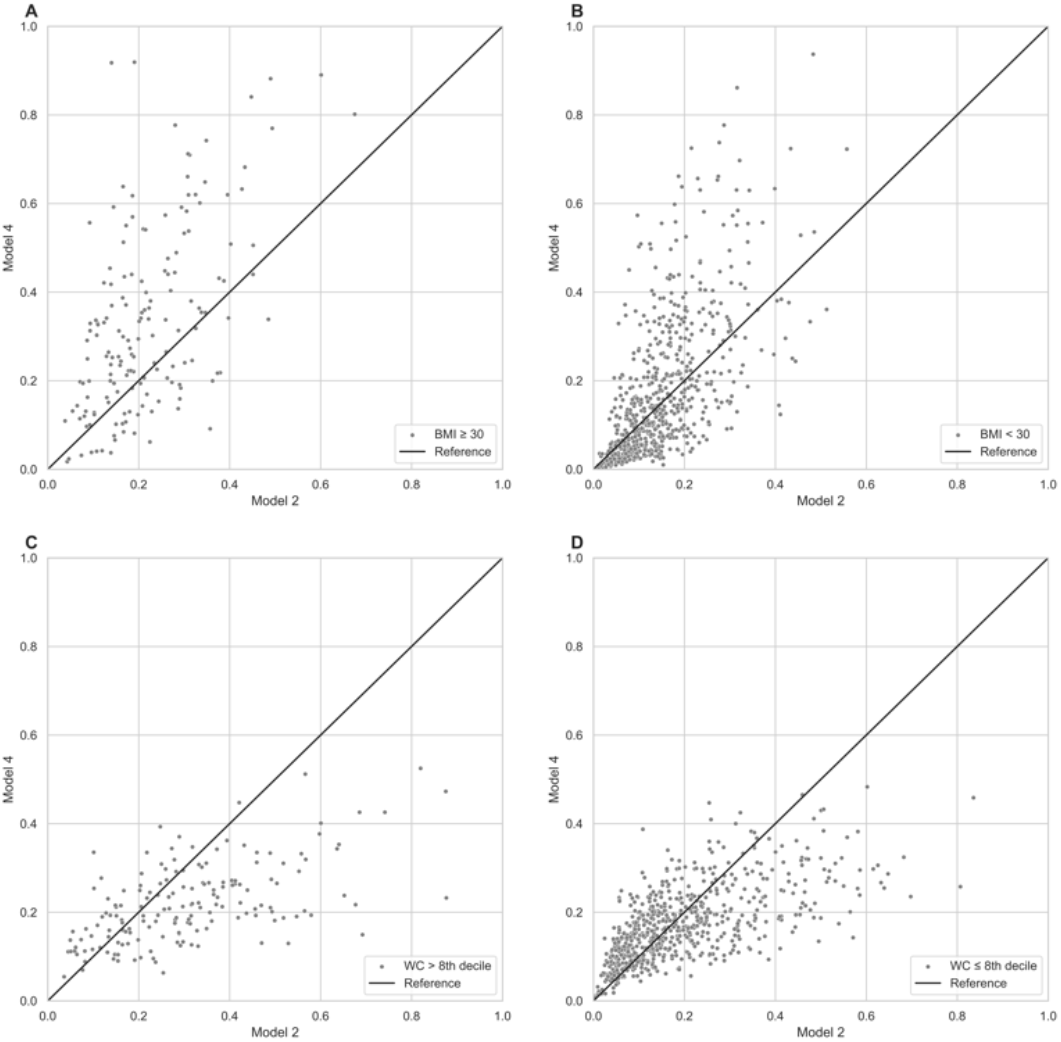
Observed and predicted means partitioned by PRS deciles. Panel A shows observed and predicted BMI means generated from Model 2 and Model 4. Similarly, Panel B shows observed and predicted waist circumference (WC) means estimated from Model 2 and Model 4.

**Table 2b** presents results of linear regression models examining the associations with offspring WC. Sex strongly associated with offspring WC and explained a substantial proportion of its variability (R^2^ = 0.19). Incorporating PRS, the change in explained WC variability was modest, yet significant, (ΔR^2^ = 0.043), and the independent contribution of sociodemographic variables was negligible (ΔR^2^=0.004; p=0.282).

**Table 2b.** Parameter Estimates from **Linear** Regression models of Waist circumference (WC) as predicted by various groups of Risk Factors.

|  |  |  |  |  |  |  |  |  |  |
| --- | --- | --- | --- | --- | --- | --- | --- | --- | --- |
|  |  | Model 1 |  | Model 2 |  | Model 3 |  | Model 4 |  |
| Group | Variable | $\beta$ | (95% CI) | $\beta$ | (95% CI) | $\beta$ | (95% CI) | $\beta$ | (95% CI) |
| 1 | Sex | 11.33*** | 9.14<br>13.52 | 11.22*** | 9.06<br>13.39 | 11.05*** | 8.89<br>13.21 | 11.29*** | 9.15<br>13.43 |
|  | PRS |  |  | 2.45*** | 1.27<br>3.64 | 2.48*** | 1.31<br>3.65 | 2.14*** | 0.90<br>3.38 |
|  | PC1 |  |  | 1.75** | 0.54<br>2.97 | 2.15** | 0.82<br>3.48 | 1.86** | 0.51<br>3.21 |
| 2 | PC2 |  |  | -0.08 | -1.16<br>1.00 | -0.23 | -1.36<br>0.89 | -0.41 | -1.49<br>0.67 |
|  | PC3 |  |  | -0.1 | -1.29<br>1.10 | -0.03 | -1.23<br>1.17 | -0.04 | -1.22<br>1.15 |
|  | PC4 |  |  | 0.47 | -1.29<br>1.10 | 0.58 | -0.68<br>1.83 | 0.61 | -0.60<br>1.82 |
|  | PC5 |  |  | 0.81* | 0.01<br>1.62 | 0.82* | -0.01<br>1.65 | 0.57 | -0.31<br>1.46 |
|  | Paternal SES |  |  |  |  | 0.4 | -0.28<br>1.08 | 0.21 | -0.46<br>0.88 |
| 3 | Maternal age |  |  |  |  | -0.15 | -0.49<br>0.18 | -0.19 | -0.51<br>0.14 |
|  | Paternal age |  |  |  |  | 0.06 | -0.23<br>0.36 | 0.04 | -0.26<br>0.33 |
|  | Birth weight |  |  |  |  |  |  | 1.4 | -0.27<br>3.07 |
|  | Gestational week |  |  |  |  |  |  | -0.66 | -1.50<br>0.17 |
| 4 | ppmBMI |  |  |  |  |  |  | 0.79*** | 0.52<br>1.07 |
|  | Parental smoking |  |  |  |  |  |  | 2.33* | 0.11<br>4.54 |
|  | GWG |  |  |  |  |  |  | 0.03 | -0.22<br>0.29 |
|  | R<sup>2</sup> | 0.1982 |  | 0.2412 |  | 0.2452 |  | 0.2877 |  |

PRS - polygenic risk score, PC – principal component SES – Socioeconomic status; ppmBMI – pre-pregnancy maternal BMI; GWG – gestational weight gain; \* p≤0.05; \*\* p≤0.01; \*\*\* p≤0.001

Incorporating perinatal variables in Model 4 (**Table 2b**), significantly increased WC explained variance (ΔR^2^= 0.0425, F_5, 823_ = 9.821; p<= 0.001). Maternal smoking and mppBMI, adjusted for other perinatal and socio-demographic characteristics and for genetic factors, were positively associated with offspring WC. The association between PRS and WC was attenuated (though remained significant) when perinatal variables were included in the model. Similar to BMI, the association between PRS deciles and WC mean levels is clearly demonstrated across the lowest and the highest deciles of PRS (**Figure 1b)**.

Additionally, logistic regression models were used to examine these associations with the dichotomous measures of obesity. Supplemental **Table S1a** reveals a significant PRS-obesity association (adjusted odds ratio [OR] per SD increment, 2.08; 95% CI, 1.56-2.77). In Model 4, OR changes for genetic variables were minor. Yet, both ppmBMI (adjusted OR per SD increment, 1.22; 95% CI, 1.14-1.30) and GWG (adjusted OR per SD increment, 1.08; 95% CI, 1.01-1.15) were significantly associated with obesity. Finally, logistic regression models applied to categorical measures of central obesity (WC > 8^th^ decile), confirmed that similar to the association with obesity, both PRS and ppmBMI’s were significantly associated with central obesity (**Table S1b**, Model 4).

**Figure 2** presents AUCs that estimate the discrimination between subjects with and without obesity (BMI ≥ 30) and high central obesity (WC > 8th decile) across the combination of risk predictors. The 10-fold average AUC increased from 0.688 (Model 2) to 0.73 (Model 4) for obesity, and from 0.65 (Model 2) to 0.68 (Model 4) for high central obesity (**Figure 2a**).

**Figure 2.**
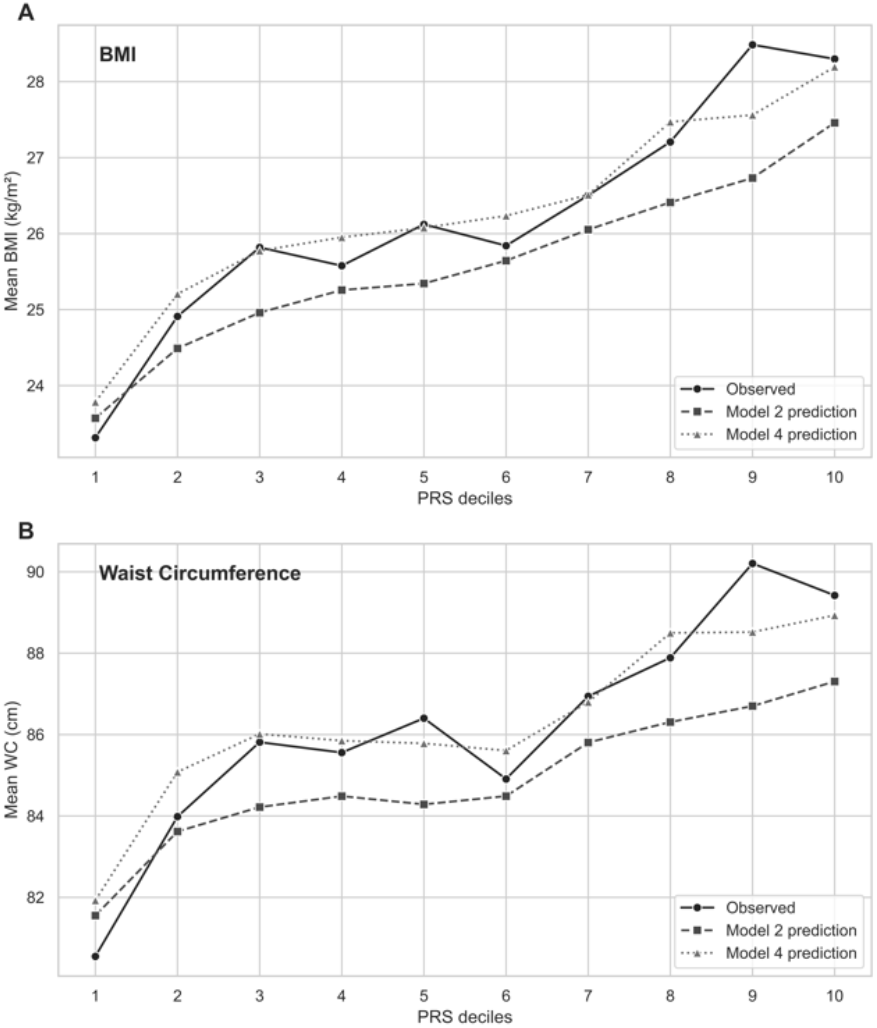
Discrimination between subjects with and without obesity, estimated by parameters of Model 2 and Model 4. Mean(±SD) AUCs are based on 10-fold cross-validation for obesity (BMI ≥ 30kg/m^2^) (Panel A) and high central obesity (WC > 8^th^ decile) (Panel A).

Next, we determined calibration by comparing observed and expected frequency of obesity-related outcomes from models without (Model 2) and with (Model 4) socio-demographic and perinatal variables. Model 2 categorized 43.9% of the sample as low risk (predicted risk ≤15%) and 56.1% as high risk (predicted risk >15%) (**Table 3**). Observed proportion of subjects without and with obesity in these 2 risk-groups were were 91.0% (negative predictive value) and 27.2% (positive predictive value), respectively. Adding socio-demographic and perinatal variables to the model (Model 4), adjusted this to 51.4% and 48.6 as low and high-risk subjects, with improved accuracy (observed proportion of subjects without and with obesity of 94.0% and 33.1%, respectively). Adding the socio-demographic and perinatal variables to the risk categories significantly improved reclassification accuracy in obesity (NRI, 0.147; 95% CI, 0.068 to 0.225), where net proportion of correct reclassification for subjects with obesity was 7/161 = 0.044 and 70/678 = 0.103 in subjects without obesity.

**Table 3.** Risk of obesity Predicted by Logistic Models with and without Perinatal characteristics. The Jerusalem Perinatal Study.

|  |  |  |  |  |  |  |  |
| --- | --- | --- | --- | --- | --- | --- | --- |
| Obesity | Predicted Risk by Model with Prenatal variables |  |  |  |  |  |  |
| | Predicted Risk by Model without Prenatal variables | <15% | $\geq 15\%$ | Overall | Reclassified as Higher Risk | Reclassified as Lower Risk | |
|  | < 15% |  |  |  |  |  |  |
|  | No. of subjects with obesity | 14 | 19 | 33 | 19 (True) | NA |  |
|  | No. of subjects without obesity | 282 | 53 | 335 | 53 (False) | NA |  |
|  | Total No. of subjects | 296 | 72 | 368 |  |  |  |
| | $\geq 15\%$ | | | | | | |
|  | No. of subjects with obesity | 12 | 116 | 128 | NA | 12 (False) |  |
|  | No. of subjects without obesity | 123 | 220 | 343 | NA | 123 (True) |  |
|  | Total No. of subjects | 135 | 336 | 471 |  |  |  |
|  | Overall |  |  |  |  |  |  |
|  | No. of subjects with obesity | 26 | 135 | 161 |  |  |  |
|  | No. of subjects without obesity | 405 | 273 | 678 |  |  |  |
|  | Total No. of subjects | 431 | 408 | 839 |  |  |  |
| Central Obesity |  |  |  |  |  |  |  |
|  | < 15% |  |  |  |  |  |  |
|  | No. of subjects with obesity | 19 | 9 | 28 | 9 (True) | NA |  |
|  | No. of subjects without obesity | 235 | 47 | 282 | 47 (False) | NA |  |
|  | Total No. of subjects | 254 | 56 | 310 |  |  |  |
| | $\geq 15\%$ | | | | | | |
|  | No. of subjects with obesity | 10 | 128 | 138 | NA | 10 (False) |  |
|  | No. of subjects without obesity | 116 | 275 | 391 | NA | 116 (True) |  |
|  | Total No. of subjects | 126 | 385 | 529 |  |  |  |
|  | Overall |  |  |  |  |  |  |
|  | No. of subjects with obesity | 29 | 137 | 166 |  |  |  |
|  | No. of subjects without obesity | 351 | 322 | 673 |  |  |  |
|  | Total No. of subjects | 380 | 459 | 839 |  |  |  |

**Figures 3a, 3b** depict predicted probabilities based on Model 4 versus Model 2 for subjects with and without obesity. We can see that, 72.7% (117/161) among subjects with obesity (BMI ≥30) had a predicted risk based on model 4 higher than the predicted risk based on model 2), i.e., a better risk estimate. Conversely, 27.3% had a higher predicted risk based on model 2. The net proportion with better risk estimates (obese NRI (>0)) hence was 45.4%. In addition, we note that 54.9% of subjects without obesity had a lower prediction based on model 4, while 45.1% had a higher prediction based on Model 4 (non-obese NRI (>0) = 9.5%). The sum of the two components of NRI (>0) equals 54.8%. The integrated discrimination improvement (IDI) also showed a significant improvement favoring the model with PRS and perinatal variables as compared to the model without perinatal characteristics (0.062%; 95%CI 0.040 to 0.083; **Table 4**)

**Table 4.** Estimates (95% CI) of Discrimination and Reclassification in Obesity-Related Variables.

|  |  |  |
| --- | --- | --- |
|  | Prediction Improvement (95% CI) |  |
| Variable | Model - 2 | Model - 4 |
|  | Sex + PCc + PRS | Sex + PCc + PRS +<br>Socidemographic + Perinatal variables |
| Obesity - C statistic | 0.701 (0.657 0.744) | 0.753 (0.714 0.793) |
| Central obesity - C statistic | 0.659 (0.615 0.704) | 0.713 (0.670 0.756) |
| Obesity – NRI* |  | 0.147 (0.068 0.225) |
| Obesity – IDI** |  | 0.062 (0.040 0.083) |
| Central Obesity – NRI* |  | 0.097 (0.034 0.159) |
| Central Obesity – IDI** |  | 0.049 (0.032 0.067) |
| * Net Reclassification index * Integrated discrimination improvement |  |  |

**Figure 3.**
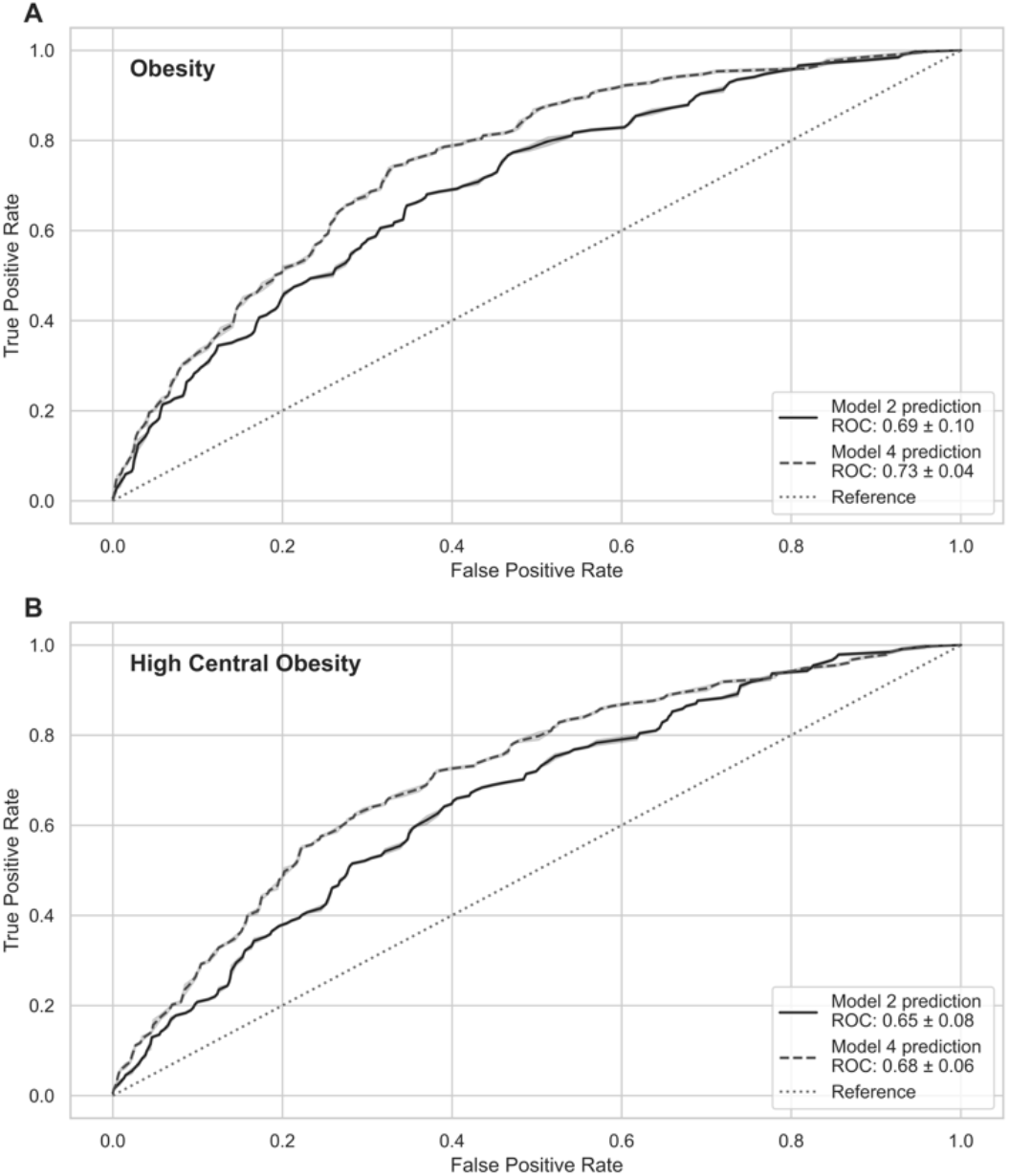
Predicted probabilities derived from Model 2 and Model 4, separately for subjects with obesity (Panel A), subjects without obesity (Panel B), subjects with high central obesity (Panel C) and without (Panel D).

Regarding central obesity, Model 2 categorized 36.9% of the sample as low risk and 63.1% as high-risk (**Table 3**). In these groups, 91.0% without and 26.1% with central obesity were observed. Model 4, compared to Model 2, placed more individuals in low risk (45.3%) and fewer in high risk (54.7%) for central obesity. The observed proportion of subjects without and with central obesity in these 2 groups were 92.4% and 29.8%, respectively. Analyses suggested somewhat better reclassification of central obesity using Model 4 (NRI, 0.097; 95% CI, 0.034 to 0.159), with net proportion of correct reclassification of -0.006 in subjects with central obesity and 0.097 in subjects without central obesity.

Similarly, **Figures 3c** and **3d** indicated that the sum of the two components of the continuous NRI (NRI>0) was about 40.0%. Yet, the better risk prediction based on Model 4, was limited to subjects with central obesity. The integrated discrimination improvement was also slightly improved (IDI=0.049; CI 0.032 to 0.067), favoring the combination of PRS and Perinatal model (**Table 4**).

## Discussion

Our association analyses revealed significant findings regarding the relationship between genetic factors and obesity-related traits in young adults. PRS displayed a significant association with these traits, albeit contributing modestly to the explained variability (4%-9%). Independent of the genetic effect, maternal characteristics during pregnancy also showed significant associations with offspring obesity-related traits. This study also demonstrated that incorporating perinatal variables into genetic risk prediction models notably improved the discrimination and reclassification of obesity risk among young adults. The results for the risk of central obesity mirrored those of overall obesity, yet the models provided slightly less accurate estimates compared to models based on BMI.

Numerous studies have detected evidence for genetic effects on obesity and related phenotypes (7). The number of susceptibility loci of obesity has grown considerably, reaching ∼200 loci for BMI (27), with most loci identified and confirmed in European populations. Given that GWAS data typically explain a small portion of phenotypic variance (27), recent studies employ PRSs to aggregate risk alleles for specific traits, aiming to gauge overall genetic susceptibility (13). Diverse obesity-related PRSs have been constructed for adults and children of different ethnic backgrounds, exhibiting significant links to obesity-related traits (12, 14, 15). For example, a large study involving middle-aged subjects (n=288,016, mean age = 57) from the UK Biobank, demonstrated a PRS approximating a normal distribution and significantly associated with BMI values (r=0.29) (15). In another large study based on young women from the Nurses’ Health Study and young men from the Health Professionals Follow-Up Study, each 10-allele increment in BMI-PRS was associated with an average BMI gain of 0.54 kg/m^2^ in women and 0.20 kg/m^2^ in men (33). In a prospective longitudinal study of participants in the Dunedin birth cohort, children with higher PRS had higher BMIs at every age assessed, from age 3 through age 38 years (14). At ages 30-38 the PRS correlation with BMI (r=0.13-0.14) and with WC (r=0.11-0.12) were statistically significant and similar.

In our study, we measured the PRS for BMI using multiple loci from SNPs derived from GWASs of BMI in the UK Biobank (34). Our analyses revealed that the correlation between PRS for BMI (with sex and 5 PCs included in the model) and BMI levels was similar (r=0.30) to that exhibited in UK biobank study, and higher than the correlation presented in the Dunedin follow-up study. The PRS correlation with WC displayed slightly lower value (r=0.20) compared to the correlation observed for BMI.

Numerous studies have examined the independent associations of obesity-PRS and early life exposure with offspring adiposity during adulthood. Our results showed independent associations between PRS and prenatal exposures like maternal excess weight during pregnancy and offspring BMI and central obesity. For instance, a one SD change in PRS correspond to 1.41 kg/m^2^ rise in offspring BMI and a 2.48 cm increase in WC. The association between PRS and offspring BMI was marginally attenuated (14.2%) with the inclusion of perinatal variables. In the WC model, a comparable modest reduction of 13.7% was observed in the PRS coefficient. It is plausible that common genetic variation contributed to both maternal and offspring phenotype levels, leading to a slight attenuation in the association between PRS and offspring obesity-related traits when perinatal variables were added to the model.

Within the Dunedin birth cohort, PRS was associated with offspring BMI independently of family history of obesity in their parents (14). Thus, PRS may contain additional information about children’s risk for obesity beyond their parental history of obesity. However, while the PRS was associated with offspring obesity at young adulthood independently of birth weight, adiposity rebound tended to mediate a large portion of PRS effect on young adulthood obesity (14).

Parental ages and socioeconomic status (SES) determined by parents’ occupation were not associated with offspring’s BMI at age 32 and their inclusion in the model didn’t change the regression coefficient for PRS. In a study spanning three generations in Finland, early-life social disadvantages related to later-life BMI, but this association weakened when accounting for PRS (35). Interestedly, in the youngest cohort included in the study, the association between early social disadvantage and later life BMI was attenuated after adjusting for PRS. The authors have suggested genetic selection and assortative mating may partly explain this social selection (35).

In another birth cohort of White Western European subjects, both lower SES and high PRS were associated with increased pre-adolescence BMI (36). The effect of SES was largely independent of the effect of the PRS and the inclusion of both variables in the model did not substantially alter the effect sizes observed when each variable was included alone.

In the second part of our analysis, we evaluated if adding early-life exposure improved the PRS prediction model’s performance by applying a 15% threshold, above which offspring would be considered at risk for obesity. Since no definitive methods for this selection is available from the literature, we were guided by the sensitivity, specificity, predictive value of a positive value (PPV), and predictive value of a negative value (NPV) as well as the number of individuals classified as high risk based on the selected threshold level. Our data showed that a risk model based on PRS had a sensitivity of 0.80 and specificity of 0.54 to predict young adult obesity (BMI ≥ 30) and the PPV and NPV of PRS were 0.27 and 0.91, respectively. Overall, the potential discrimination for predicting offspring’s obesity increased when the perinatal variables were included in the model (sensitivity = 0.84, specificity = 0.60, PPV = 0.33 and NPV = 0.94). Our analysis also indicated that the PRS model’s ability to discriminate between individuals with and without obesity (AUC= 0.69) was improved when perinatal variables were included (AUC = 0.73).

Several studies have examined the obesity risk prediction models based on PRS. Despite the difference in the number of loci, the models’ ability to discriminate between individuals with and without obesity did not substantially improved (AUC ≤0.61)(7). Two studies, one that used a model which included the 12 most strongly BMI-associated loci, including FTO and MC4R (37) and another study where the used model included additional 20 loci (38) showed exactly the same prediction of overall performance (AUC of 0.57).

In our study, in addition to PRS, maternal pre-pregnancy BMI provided a major contribution to the improvement in the model ability to discriminate between individuals with and without obesity. Another study has shown that parental obesity status at age 6–9 years had a sensitivity, specificity, PPV and NPV of 0.8, 0.70, 0.27 and 0.90 respectively, to predict adult obesity (7, 39). This may suggest that family history may combine both genetic and non-genetic components which contribute to the obesity prediction in offspring. It has been shown that the sensitivity of parental obesity for predicting the offspring’s adult obesity risk may increase with the age at which parental obesity is assessed, whereas the specificity decreases with age. This can partially explain the higher sensitivity and lower specificity obtained in our study as compared to the findings described above, due to the fact that in our study pre-pregnancy maternal BMI was determined at older ages. In a Danish study, the AUC for a genetic model (PRS only) was 0.58 and after the inclusion of non-genetic obesity risk factors (e.g. education, diet, smoking, and physical activity) the AUC increased to 0.69 (40).

While inclusion of perinatal variables to the PRS prediction model resulted in numerous changes in offspring obesity risk, only small proportion of the total JPS study sample (8.6%), was reclassified as high risk and 16.1% were reclassified as having low risk. While, 8.7% of the obesity events (14/161) occurred among individuals who were not classified as high risk either by PRS alone or by PRS and perinatal exposures, 72% of the events (116/161) occurred among individuals who were classified as high risk both by PRS alone and by the PRS and perinatal risk combined model.

Inspection of the relative contribution of correct reclassification for events and non-events revealed important strengths and weaknesses of the combined PRS-perinatal strategy. For the entire cohort, the NRI for obesity-events was 0.147, whereas the NRI for non-events was 0.103 and for events 0.044. For our study subjects the PRS and Perinatal obesity model (at 15% risk threshold), the high NPV (0.94) provides confidence that a limited number of offspring who were identified as low risk will become overweight or obese at age 30 and therefore may miss out a targeted program aimed to lower their weight. However, a significant proportion of children identified as high risk, (including those who were reclassified as being high risk) will obviously not become obese by age 30 years.

The advantage of the PRS and early life exposures is that they can be assessed at young ages long time before the adverse outcomes are initiated. Using these markers for early screening may identify high-risk individuals who could truly benefit from early behavioral programs for lifestyle modification (e.g. health diet and increased physical activity). The potential harms of such behavioral and/or environmental program to tackle obesity, if applied also to low-risk individuals, are likely to be low.

The major strength of our study is the combination of high-quality detailed records of prenatal and perinatal characteristics with a comprehensive long-term follow-up data.

There are several limitations to our study. First, it includes only a sample of offspring from the original 1974-1976 JPS cohort who were invited to participate in the follow-up study. However, using a stratified sampling approach and over-sampling in the ends of the distribution ensured that offspring with a full range of mppBMI and birth weight were included in our study. Second, while the PRS explained only a modest percent of the variation for BMI, this is similar to other studies and expected with the current methodology used (15, 35). Third, both mppBMI and GWG were reported by mothers in interviews conducted by nurses while hospitalized after delivery. Verification from clinical records was not available. Nevertheless, we have demonstrated associations between reported maternal attributes and BMI more than 30 years later, as well as with long-term clinical outcomes in mothers (41), which lend support to the validity of the data. In our study, mothers were interviewed within days after delivery, in accordance of the reports on the validity and reproducibility of maternal recollection of pre-pregnancy weight (42, 43). Even if reporting error was present it most likely that this non-differential misclassification resulted in an underestimation in our findings.

This study points to the strong relationship between BMI and obesity risk during early adulthood with PRS and with maternal excess weight before pregnancy. In addition, our prediction models demonstrate that utilizing PRS plus prenatal exposures enhances the ability to quantify future obesity risk in young adults and thus form the basis for early term preventive programs.

## Data Availability

The JPS cohorts was introduced in 1976 and research using this cohort were published along the last 2 decades.

## Supplementary Table S1a, S1b

**Table S1a.** Parameter Estimates from **Logistic** models of **Obesity** as predicted by various Groups of Risk Factors.

|  |  |  |  |  |  |  |  |  |  |
| --- | --- | --- | --- | --- | --- | --- | --- | --- | --- |
|  |  | Model 1 |  | Model 2 |  | Model 3 |  | Model 4 |  |
| Group | Variable | OR | (95% CI) | OR | (95% CI) | OR | (95% CI) | OR | (95% CI) |
| 1 | Sex | 1.28 | 0.76 - 2.17 | 1.30 | 0.74 - 2.27 | 1.22 | 0.69 - 2.17 | 1.21 | 0.66 - 2.24 |
|  | PRS |  |  | 2.08*** | 1.56 - 2.77 | 2.11*** | 1.57 - 2.83 | 1.99*** | 1.43 - 2.76 |
|  | PC1 |  |  | 1.56** | 1.14 - 2.13 | 1.87*** | 1.28 - 2.72 | 1.84** | 1.21 - 2.78 |
| 2 | PC2 |  |  | 1.06 | 0.80 - 1.40 | 0.98 | 0.72 - 1.33 | 0.92 | 0.67 - 1.26 |
|  | PC3 |  |  | 0.93 | 0.68 - 1.26 | 0.94 | 0.68 - 1.30 | 0.97 | 0.69 - 1.36 |
|  | PC4 |  |  | 1.21 | 0.89 - 1.66 | 1.26 | 0.91 - 1.77 | 1.34 | 0.97 - 1.84 |
|  | PC5 |  |  | 1.39* | 1.04 - 1.85 | 1.37* | 1.03 - 1.82 | 1.39* | 1.02 - 1.88 |
|  | Paternal SES |  |  |  |  | 1.19 | 0.99 - 1.44 | 1.13 | 0.93 - 1.37 |
| 3 | Maternal age |  |  |  |  | 0.96 | 0.88 - 1.05 | 0.95 | 0.86 - 1.04 |
|  | Paternal age |  |  |  |  | 0.98 | 0.90 - 1.06 | 0.97 | 0.89 - 1.06 |
|  | Birth weight |  |  |  |  |  |  | 0.86 | 0.55 - 1.36 |
|  | Gestational week |  |  |  |  |  |  | 0.93 | 0.75 - 1.16 |
| 4 | ppmBMI |  |  |  |  |  |  | 1.22*** | 1.14 - 1.30 |
|  | Parental smoking |  |  |  |  |  |  | 1.59 | 0.86 -2.95 |
|  | GWG |  |  |  |  |  |  | 1.08* | 1.01 - 1.15 |
|  | -2lnL | 2,270.10 |  | 2,097.13 |  | 2,045.97 |  | 1,899.76 |  |
| PRS - polygenic risk score, PC – principal component SES – Socioeconomic status; ppmBMI – pre-pregnancy maternal BMI; GWG – gestational weight gain; * p≤0.05; ** p≤0.01; *** p≤0.001 |  |  |  |  |  |  |  |  |  |

**Table S1b.** Parameter Estimates from **Logistic** models of **Central Obesity** as predicted by various Groups of Risk Factors.

|  |  |  |  |  |  |  |  |  |  |
| --- | --- | --- | --- | --- | --- | --- | --- | --- | --- |
|  |  | Model 1 |  | Model 2 |  | Model 3 |  | Model 4 |  |
| Group | Variable | OR | (95% CI) | OR | (95% CI) | OR | (95% CI) | OR | (95% CI) |
| 1 | Sex | 1.3 | 0.78 - 2.16 | 1.29 | 0.76-2.19 | 1.2 | 0.69-2.08 | 1.24 | 0.70-2.21 |
|  | PRS |  |  | 1.67*** | 1.29-2.15 | 1.67*** | 1.30-2.16 | 1.63*** | 1.24-2.15 |
|  | PC1 |  |  | 1.5** | 1.11-2.03 | 1.72** | 1.21-2.45 | 1.66** | 1.15-2.40 |
| 2 | PC2 |  |  | 1.11 | 0.84-1.45 | 1.04 | 0.79-1.37 | 1.01 | 0.77-1.33 |
|  | PC3 |  |  | 0.99 | 0.75-1.30 | 1.01 | 0.75-1.35 | 1 | 0.74-1.35 |
|  | PC4 |  |  | 1.26 | 0.95-1.67 | 1.31 | 0.97-1.76 | 1.33 | 0.99-1.80 |
|  | PC5 |  |  | 1.44* | 0.95-1.67 | 1.43* | 1.07-1.91 | 1.42* | 1.05-1.92 |
|  | Paternal SES |  |  |  |  | 1.12 | 0.96-1.30 | 1.07 | 0.92-1.24 |
| 3 | Maternal age |  |  |  |  | 0.94 | 0.85-1.05 | 0.94 | 0.85-1.04 |
|  | Paternal age |  |  |  |  | 1 | 0.91-1.10 | 0.99 | 0.90-1.09 |
|  | Birth weight |  |  |  |  |  |  | 1.21 | 0.78-1.86 |
|  | Gestational week |  |  |  |  |  |  | 0.88 | 0.71-1.08 |
| 4 | ppmBMI |  |  |  |  |  |  | 1.16*** | 1.08-1.23 |
|  | Parental smoking |  |  |  |  |  |  | 1.75 | 0.97-3.14 |
|  | GWG |  |  |  |  |  |  | 0.99 | 0.93-1.06 |
|  | -2lnL | 2,429.21 |  | 2,299.87 |  | 2,260.80 |  | 2,160.59 |  |
| PRS - polygenic risk score, PC – principal component SES – Socioeconomic status; ppmBMI – pre-pregnancy maternal BMI; GWG – gestational weight gain; * p≤0.05; ** p≤0.01; *** p≤0.001 |  |  |  |  |  |  |  |  |  |

